# A common allele of HLA mediates asymptomatic SARS-CoV-2 infection

**DOI:** 10.1101/2021.05.13.21257065

**Authors:** Danillo G. Augusto, Tasneem Yusufali, Joseph J. Sabatino, Noah D. Peyser, Lawton D. Murdolo, Xochitl Butcher, Victoria Murray, Vivian Pae, Sannidhi Sarvadhavabhatla, Fiona Beltran, Gurjot Gill, Kara Lynch, Cassandra Yun, Colin Maguire, Michael J. Peluso, Rebecca Hoh, Timothy J. Henrich, Steven G. Deeks, Michelle Davidson, Scott Lu, Sarah A. Goldberg, J. Daniel Kelly, Jeffrey N. Martin, Cynthia A. Viera-Green, Stephen R. Spellman, David J. Langton, Sulggi Lee, Gregory M. Marcus, Jeffrey E. Olgin, Mark J. Pletcher, Stephanie Gras, Martin Maiers, Jill A. Hollenbach

## Abstract

Despite some inconsistent reporting of symptoms, studies have demonstrated that at least 20% of individuals infected with severe acute respiratory syndrome coronavirus 2 (SARS-CoV-2) will remain asymptomatic. Although most global efforts have focused on understanding factors underlying severe illness in COVID-19 (coronavirus disease of 2019), the examination of asymptomatic infection provides a unique opportunity to consider early disease and immunologic features promoting rapid viral clearance. Owing to its critical role in the immune response, we postulated that variation in the human leukocyte antigen (HLA) loci may underly processes mediating asymptomatic infection. We enrolled 29,947 individuals registered in the National Marrow Donor Program for whom high-resolution HLA genotyping data were available in the UCSF Citizen Science smartphone-based study designed to track COVID-19 symptoms and outcomes. Our discovery cohort (n=1428) was comprised of unvaccinated, self-identified subjects who reported a positive test result for SARS-CoV-2. We tested for association of five HLA loci (HLA-A, -B, -C, -DRB1, -DQB1) with disease course and identified a strong association of HLA-B*15:01 with asymptomatic infection, and reproduced this association in two independent cohorts. Suggesting that this genetic association is due to pre-existing T-cell immunity, we show that T cells from pre-pandemic individuals carrying HLA-B*15:01 were reactive to the immunodominant SARS-CoV-2 S-derived peptide NQKLIANQF, and 100% of the reactive cells displayed memory phenotype. Finally, we characterize the protein structure of HLA-B*15:01-peptide complexes, demonstrating that the NQKLIANQF peptide from SARS-CoV-2, and the highly homologous NQKLIANAF from seasonal coronaviruses OC43-CoV and HKU1-CoV, share similar ability to be stabilized and presented by HLA-B*15:01, providing the molecular basis for T-cell cross-reactivity and HLA-B*15:01-mediated pre-existing immunity.

## Introduction

Despite some inconsistent reporting of symptoms,^1^ studies have demonstrated that at least 20% of individuals infected with severe acute respiratory syndrome coronavirus 2 (SARS-CoV-2) will remain asymptomatic.^2–4^ Although most global efforts have focused on understanding factors underlying severe illness in COVID-19 (coronavirus disease of 2019), the examination of asymptomatic infection provides a unique opportunity to consider early disease and immunologic features promoting rapid viral clearance. Specific focus on asymptomatic infection has the potential to further our understanding of disease pathogenesis and supports ongoing efforts toward vaccine development and the discovery of potential therapeutic targets.

It remains unclear why many individuals successfully clear infection without major complications while others develop severe disease, even in the absence of known risk factors for severe COVID-19 outcomes.^5^ However, host genetics is long-known to be implicated in differential immunological responses to infection and disease progression.^6^ Numerous studies intending to understand the genetic basis of differential outcomes in COVID-19 have been underway since nearly the start of the global pandemic, including the multicenter Host Genetics Initiative.^7^ However, the vast majority of these studies have examined genetic associations with severe disease course, in primarily hospitalized cohorts.^8–10^ As a result, although most individuals infected with SARS-CoV-2 experience mild disease course or are entirely asymptomatic, very few studies have examined genetics in the context of non-hospitalized, prospective, community-based cohorts.

The *human leukocyte antigen (HLA)* region, located on chromosome 6p21, is the most polymorphic and medically important region of the human genome. Variation in *HLA* has been associated with hundreds of diseases and conditions, including infection. Infectious diseases are one of the leading causes of human mortality^11^ and are a primary selective pressure shaping the human genome, and specifically *HLA*.^12,13^ Among the myriad genes involved in human immune responses, *HLA* variants are among the strongest reported associations with viral infections. For example, *HLA* was strongly associated with the rapid progression and viral load control of HIV (human immunodeficiency virus),^14^ hepatitis B, hepatitis C, and other infectious diseases.^15^ Notably, *HLA* class I and class II alleles have also been associated with the severe acute respiratory syndrome caused by SARS-CoV.^16–21^

A series of *in silico* analyses have pointed to HLA as relevant molecules for SARS-CoV-2 risk and essential targets for vaccine development.^22–25^ For example, *HLA-B*46:01* has low predicted binding of peptides for SARS-CoV-2, suggesting that individuals expressing this molecule may be more vulnerable to COVID-19,^24^ which corroborates previous results showing *HLA-B*46:01* association with SARS risk.^18^ In contrast, *HLA-B*15:03* was predicted to protect against COVID-19 with the greatest ability to present highly conserved SARS-CoV-2 peptides to T cells.^24^ More recently, it was demonstrated that while there is some overlap, many SARS-CoV-2 epitopes for CD8+ T cells are *HLA* specific.^24^ To date, relatively few studies have directly examined HLA associations with infection, with mixed and inconclusive results in relatively small cohorts.^26^ Larger studies that relied on genome-wide data to impute *HLA* failed to find robust associations with disease;^8,27^ however, these studies focused primarily on hospitalized patients with severe disease course.

Because of its pivotal role in the immune response, understanding the impact of *HLA* variation in disease promises to provide meaningful insights relevant to understanding the immunopathogenesis of COVID-19, while informing vaccine development and potential immunotherapies. Here, we present the largest study to-date directly examining *HLA* variation in the context of primarily mild disease. We invited volunteer bone marrow donors for whom high-resolution *HLA* genotyping data were already available to participate in the COVID-19 Citizen Science Study, a smartphone-based study designed to track COVID-19 symptoms and outcomes, including self-reported positive tests for SARS-CoV-2 infection, to develop a prospective cohort currently numbering nearly 30,000 individuals, as well as two additional independent cohorts. We further contextualize our findings by examining T cell reactivity and structural implications for the observed HLA associations. Our results provide strong support for the role of HLA class I in viral clearance leading to asymptomatic infection among persons with SARS-CoV-2 infection and provide an important framework for additional studies aimed at revealing the immunological and genetic basis for recovery from SARS-CoV-2 infection.

## Methods

### Data collection, discovery (Citizen Science) cohort

Subjects were volunteer bone marrow donors with valid email addresses on file with the National Marrow Donor Program (NMDP) who were invited to participate in the study through an email outreach campaign that began in July 2020. All subjects have within the NMDP database a pre-existing record for high-resolution *HLA* genotyping, typically for five loci (*HLA-A, -B, -C, - DRB1*, and *-DQB1*).^28^ Participants who opt in to the study are required to download a smartphone app and participate in the COVID-19 Citizen Science Study (launched using the Eureka Digital Research Platform, https://eureka.app.link/covid19) or, as of January 2021, participate via website (https://covid19.eurekaplatform.org/). Once enrolled, participants are asked to complete an initial 10- to 15-minute survey about baseline demographics, their health history and daily habits. Follow-up daily questions specific to symptoms, weekly questions regarding testing and monthly questions regarding hospitalization for COVID-19, are delivered by push notification or text message on an ongoing basis and require five to 15 minutes per week. As of April 30, 2021, we enrolled 29,947, of whom 21,893 have completed their baseline survey. Participation in the UCSF Citizen Science study and linking to NMDP HLA data were approved by the Institutional Review Board for the University of California, San Francisco (IRB# 17-21879 and IRB# 20-30850, respectively).

Within the mobile application, survey respondents are asked during their initial baseline survey whether they have ever been tested for active infection and report the result (positive, negative, do not know) and the approximate number of weeks since the test. Thereafter, each week respondents are asked whether they were tested in the prior week, and to report the result. We considered anybody reporting a positive test for active infection as having been infected with SARS-CoV-2. Our cohort consisted of individuals reporting a positive test for virus up to April 30, 2021, prior to the implementation of widespread vaccination for the virus. We restricted the analysis to individuals who had self-identified as “White” only due to insufficient numbers for analysis in other groups, allowing analysis of 1428 individuals. Inclusion criteria are detailed in **Figure 1**.

**Figure 1.**
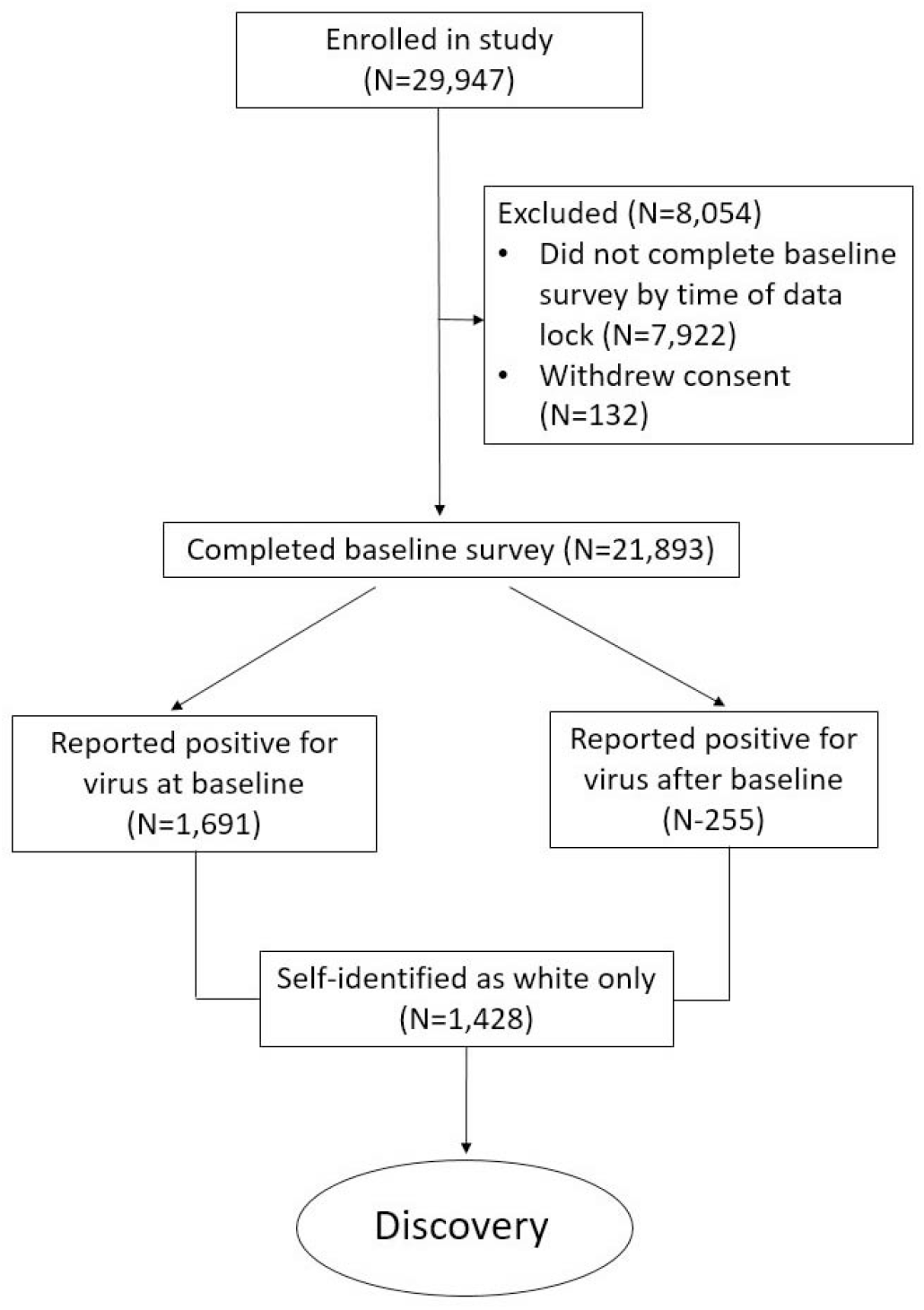
Discovery cohort inclusion criteria.

Symptoms are self-reported at baseline and in daily surveys. Within the baseline survey, respondents are asked to report whether they had any of a list of symptoms (**Supplementary Table S1**) for three days or longer at any time since February of 2020. These same symptoms are queried in each daily survey, where respondents are asked whether they experienced each symptom within the previous 24 hours. Among those individuals, we considered those who reported having had a positive test for active virus at baseline, with a time since the test of longer than two weeks or who did not specify test dates, and who reported “None of the above” for all symptoms in the baseline survey, as “asymptomatic.” We also considered daily symptom reports for the two weeks after baseline for respondents who reported a positive test for active infection at baseline as having occurred within the prior two weeks. In these cases, we considered individuals asymptomatic if in addition to reporting no symptoms at baseline, they did not report any single symptom two or more times within this time period. For individuals who did not report a positive test for active infection at baseline, but subsequently reported a positive test on a weekly survey, we used the same criteria considering daily symptom reports for the period two weeks prior and two weeks after the positive test report (**Figure 2**).

**Figure 2.**
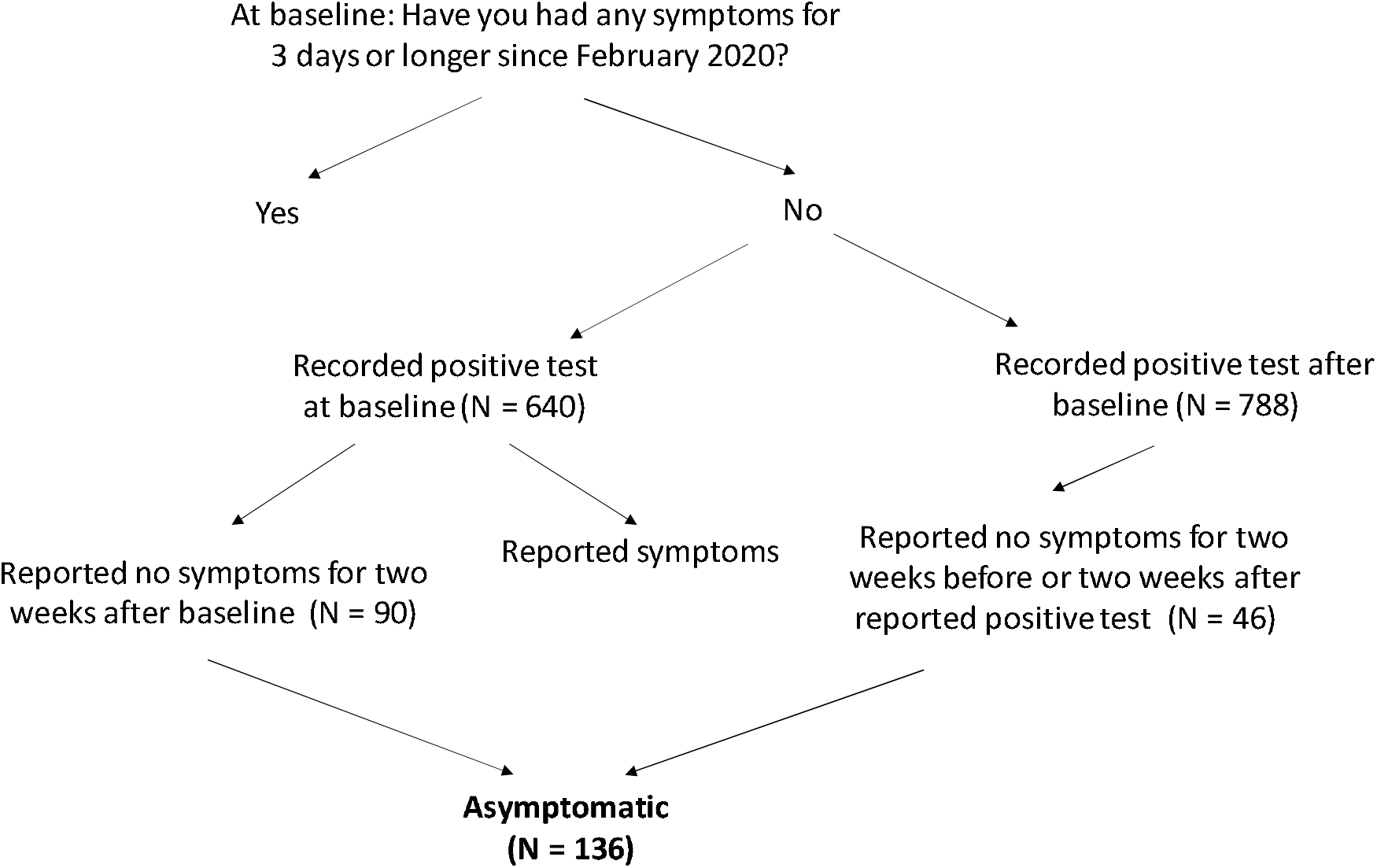
Definition of asymptomatic disease course in the discovery cohort.

### CHIRP and LIINC (replication) cohorts

Study participants were enrolled in two UCSF-based prospective longitudinal cohorts: the COVID-19 Host Immune Response Pathogenesis (CHIRP) study and the Long-term Impact of Infection with Novel Coronavirus (LIINC) study. Participants were identified through local clinical systems (UCSF Moffitt Hospital, San Francisco General Hospital, Kaiser, California Pacific Medical Center, etc.) as well as the San Francisco Department of Public Health. After confirmation of SARS-CoV-2 test results or exposure to determine eligibility, participants were asked to sign a consent form, complete a baseline visit, and schedule follow-up in-person visits. The CHIRP study included volunteers with positive PCR test documentation and/or symptom onset within the preceding 21 days. Asymptomatic disease was defined as having a confirmed positive PCR test with lack of any symptoms (“Did you have or are you still having any symptoms that you think are because of COVID-19?”) at baseline and follow-up visits. A total of five longitudinal samples were collected from acute COVID-19 infected participants. The first sample was collected <31 days of symptom onset or <31 days from exposure to SARS-CoV-2 as a week 0 baseline visit. The remaining samples were collected at weeks 1, 3, 10, and 24. At each CHIRP visit, blood and nasopharyngeal (NP) swabs were collected. Optional sample collection included sputum, saliva stool, and urine. The LIINC study enrolled participants with prior SARS-CoV-2 infection confirmed on clinical nucleic acid amplification testing between 14 and 90 days after initial symptom onset. Following written informed consent, clinical data and biospecimens were collected monthly for up to 4 months after initial symptom onset and then every 4 months thereafter. Biospecimens including blood and saliva were collected at each visit. CHIRP and LIINC utilized a harmonized set of case report forms to collect clinical data about demographics, medical history, the COVID-19 illness, and post-acute symptoms. Clinical labs collected during in-person CHIRP visits included complete blood count with differential, comprehensive metabolic panel, erythrocyte sedimentation rate (ESR), high sensitivity C-reactive protein (hs-CRP), D-dimer, lactate dehydrogenase (LDH), and ferritin. All participants provided written informed consent, and the CHIRP and LIINC studies were approved by the Institutional Review Board for the University of California, San Francisco, (IRB# 20-30588 and 20-30479, respectively).

### HLA genotyping in the CHIRP/LIINC cohort

A total of 100 ng of high-quality DNA was fragmented using the Library Preparation Enzymatic Fragmentation Kit 2.0 (Twist Bioscience, San Francisco, USA). Subsequently, the ends of the fragmented DNA were repaired, poly-A tail was added and ligated through PCR to Illumina-compatible dual index adapters that were uniquely barcoded. After ligation, fragments were purified with 0.8X ratio AMPure XP magnetic beads (Beckman Coulter, Brea, USA), followed by double-size selection (0.42X and 0.15X ratios) to select libraries of approximately 800 bp. Finally, libraries were amplified and purified with magnetic beads.

After fluorometric quantification, 30 ng of each sample was precisely pooled using ultrasonic acoustic energy, and the targeted capture was performed with Twist Target Enrichment kit (Twist Bioscience, San Francisco, USA). Briefly, the volumes were reduced using magnetic beads, and the DNA libraries were bound to 1394 biotinylated probes specific to the HLA region, covering all exons, introns, and regulatory regions of *HLA-A, HLA-B, HLA-C, HLA-DRB1, HLA-DRA, HLA-DQB1, HLA-DQA1, HLA-DPB1*, and *HLA-DPA1*. Fragments targeted by the probes were captured with streptavidin magnetic beads and then amplified and purified. Enriched libraries were analyzed in BioAnalyzer (Agilent, Santa Clara, USA) and quantified by digital-droplet PCR. Finally, enriched libraries were sequenced with the NovaSeq platform (Illumina, San Diego, USA) with a paired-end 150 bp sequencing protocol. After sequencing, data were analyzed with HLA Explorer (Omixon, Budapest, Hungary) and AlloSeq Tx (CareDx, Brisbane, USA).

### UK cohort re-analysis

We re-analyzed the primary data from the study by Langton et al. (2021)^29^ in which it was performed the analysis of HLA class I and class II genes in 147 individuals of European ancestry with known SARS-CoV-2 infection and a range of symptoms and 69 asymptomatic hospital workers. In the initial publication, *HLA-B*15:01* was not directly tested for association with asymptomatic disease course. *HLA* genotyping methods, allele frequencies, demographics, and clinical outcomes are as previously described.

### HLA association analysis

In our discovery cohort, we examined the association of five *HLA* loci (*HLA-A, -B, -C, -DRB1, - DQB1*) with asymptomatic vs. symptomatic infection. Initial testing for *HLA* associations was performed using the R package BIGDAWG^30^, which handles multiallelic *HLA* data to test for association at the haplotype, locus, allele, and amino acid levels. We next employed a generalized linear model using ‘glm’ in the R base package to consider relevant covariates, including any reported comorbidities, sex, and age, for alleles initially found to be associated with asymptomatic infection after correction for multiple testing. We corrected p-values using the Bonferroni method^31^ for the number of alleles tested at *HLA-A, -B*, and *-DRB1*, which accounts for the strong linkage disequilibrium between some loci tested. For the replication cohorts, we tested only the allele of interest, using the generalized linear model framework as described. Meta-analysis of results in our three cohorts was performed in R using the common effect model with the ‘meta’ package.^32^

### HLA-B*15:01 tetramer analysis

SARS-CoV-2 peptides were synthesized by in vitro transcription and translation (IVTT) of oligonucleotides encoding each peptide (**Supplementary Table S2**) using the PURExpress in vitro protein synthesis kit (New England BioLabs, Ipswich, USA) as previously described.^33^ Each peptide was loaded on biotinylated HLA-B*15:01 easYmers (immunAware, Copenhagen, Denmark) according to manufacturer’s instructions. Peptide-loaded HLA-B*15:01 tetramers were generated using streptavidin conjugated to phycoerythrin (PE), allophycocyanin (APC), PE-CF594, or BV421 according to the manufacturer’s instructions. 500 µM D-biotin was subsequently added to each peptide-loaded tetramer and tetramers were pooled just prior to cell staining. Combinatorial tetramer staining was used to identify each epitope by unique combinations of dual fluorophores where at least one of the fluorophores contained PE (**Supplementary Table S3**). For this experiment, we obtained peripheral blood mononuclear cells (PBMCs) stored in the National Marrow Donor Program® (NMDP)/Be The Match® Research Sample Repository (ClinicalTrials.gov protocol # NCT04920474) that had been collected from nine healthy donors prior to the start of the COVID-19 pandemic. All individuals consented to research and had been previously genotyped for *HLA* class I and class II and were determined to be homozygous for HLA-B*15:01. Demographic details and full genotyping are given in **Supplementary Table S4**. The frequencies of antigen-specific T cells were calculated as previously described.^34^ Briefly, an aliquot of PBMCs was used for cell surface staining and counting with 123count eBeads (Invitrogen, Waltham, USA). The remaining PBMCs were stained with the indicated tetramer pools (**Supplementary Table S5**) and enriched using anti-PE magnetic microbeads (Miltenyi, Bergisch Gladbach, Germany) over a magnetic column, cell-surface stained, and counted as for pre-enrichment. CD8+ T cells were identified by gating of live singlet CD8+ lymphocytes that were negative for CD4/CD14/CD16/CD19. A stringent tetramer gating strategy was employed whereby CD8+ T cells labeled with only two fluorophores were considered antigen-specific. Memory status of tetramer-positive CD8+ T cells was determined by lack of CCR7 and CD45RA co-expression (i.e. all non-naïve cells). Given the low numbers of cells available from donors, only tetramer-positive CD8+ T cells with frequencies greater than 1 × 10^−4^ per total CD8+ T cells were considered positive.

### Protein expression, refold and purification of pHLA complexes

pET-30a(+) DNA plasmids encoding HLA-B*15:01 α-chain and human β2-microglobulin were transformed separately into BL21 *E. coli* and expressed. Inclusion bodies containing the individually expressed recombinant proteins were extracted, purified, and quantified as previously described.^35^ Soluble peptide-HLA-B*15:01 complexes were produced by refolding 30 mg of HLA-B*15:01 α-chain with 10 mg of β2-microglobulin and 5 mg of peptide (Genscript, Piscataway, USA) into a buffer of 0.5 M L-Arginine (Sigma-Aldrich, St Louis, USA), 0.1 M Tris-HCl pH 8.0 (Fisher Bioreagents, Waltham, USA), 2.5 mM EDTA pH 8.0 (Sigma-Aldrich, St Louis, USA), 5 mM Glutathione (reduced) (Goldbio, St Louis, USA), 1.25 mM Glutathione (Goldbio, St Louis, USA). The peptides selected are summarized in **Table 1**. The refold mixture was dialyzed at 4°C in 10 mM Tris-HCl pH 8.0 (3 times for 12 hours) and soluble pHLA complexes were purified via anion exchange chromatography.

**Table 1.** Stability of HLA-B*15:01 in complex with the NQK peptides from SARS-CoV-2 and OC43 viruses.

| Peptide sequence | Viral origin | T<sub>m</sub> (°C) ± S.E.M. |
| --- | --- | --- |
| NQKLIAN**Q**F | SARS-CoV-2 | 51.60 ± 0.38 |
| NQKLIANA**F** | OC43-CoV and HKU1-CoV | 51.01 ± 0.11 |

The **bold** letters show the difference in sequence of SARS-CoV-2 peptides compared to OC43-CoV and HKU1-CoV. The T<sub>m</sub> designate the thermal melt temperature (n = 2).

### Differential scanning fluorimetry

Thermal stability assay was performed by Differential Scanning Fluorimetry carried out in ViiA 7 real-time PCR system (Thermofisher, Waltham, USA), where HLA-B*15:01-YFP complex was heated from 25 to 95°C at a rate of 1°C/min in 0.5°C steps. The excitation and emission channels were set to the TAMRA reporter (x3m3 filter) with excitation of ∼550 nm and detection at ∼587 nm. The experiment was performed at two concentrations of pHLA (5 µM and 10 µM) in duplicates. Each sample was dialyzed in 10mM Tris-HCl pH 8.0 (Fisher Bioreagents, Waltham, USA), 150mM NaCl and contained a final concentration of 10X SYPRO Orange Dye (Invitrogen, Waltham, USA). Fluorescence intensity data was normalized and plotted using GraphPad Prism 9 (version 9.3). The Tm value for a pHLA is equal to its the temperature when 50% of maximum fluorescence intensity is reached, which approximately equals to 50% of unfolded protein and is summarized in **Table 1**.

### Crystallization and structure determination

Crystals of HLA-B*15:01-peptide complexes were grown via hanging-drop vapor diffusion at 20°C. The protein: reservoir drop ratio was 1:1, at a concentration of 3 mg/mL in 10 mM Tris-HCl pH 8 (Fisher Bioreagents, Waltham, USA), 150 mM NaCl (Merck, Darmstadt, Germany). Crystals of HLA-B*15:01-NQKLIAN**Q**F (SARS-CoV-2) were grown in 0.2 M Sodium Formate pH 7.0 and 20% w/v Polyethylene glycol 3,350; and for HLA-B*15:01-NQKLIAN**A**F (HKU1-CoV and OC43-CoV) in 20% w/v Polyethylene glycol 3,350 and 2% v/v Ethylene glycol. Protein crystals were soaked in a cryoprotectant solution containing mother liquor solution with 20% (v/v) Ethylene glycol and then flash-frozen in liquid nitrogen. The data were collected on the MX2 beamline at the Australian Synchrotron, part of ANSTO, Australia.^36^ The data were processed using XDS^37^ and the structures were determined by molecular replacement using the PHASER program^38^ from the CCP4 suite^39^ with a model of HLA-B*15:01 without the peptide (derived from PDB ID: 5TXS).^40^ Manual model building was conducted using COOT^41^ followed by refinement with BUSTER^42^ and PHENIX.^43^ The final models have been validated and deposited using the wwPDB OneDep System and the final refinement statistics, and PDB codes are summarized in **Table 2**. All molecular graphics representations were created using PyMOL.

**Table 2.**
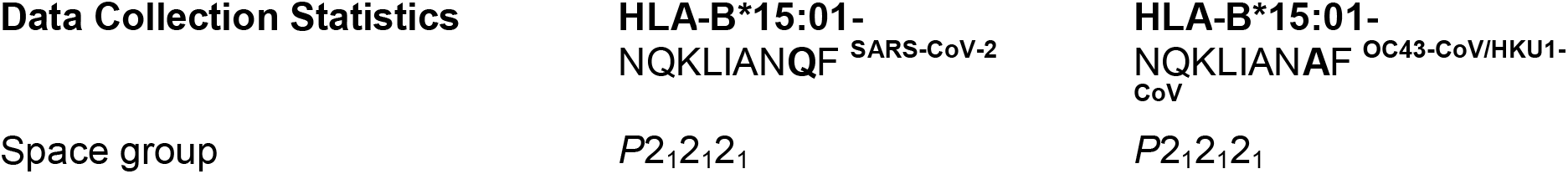
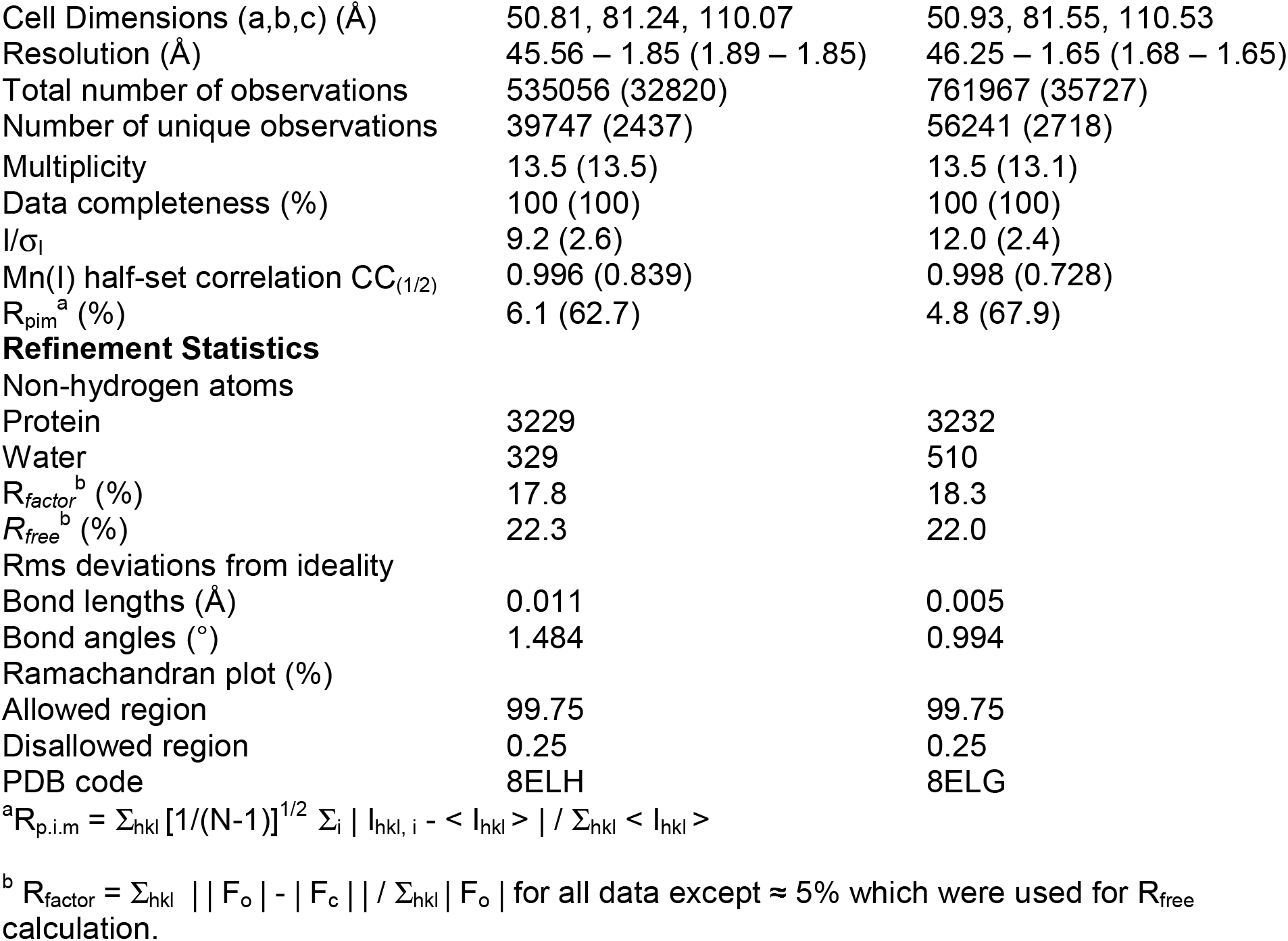
Data Collection and Refinement Statistics.

| **Data Collection Statistics** | **HLA-B\*15:01-**<br>**NQKLIANQF<sup>SARS-CoV-2</sup>** | **HLA-B\*15:01-**<br>**NQKLIANAF<sup>OC43-CoV/HKU1-CoV</sup>** |
| --- | --- | --- |
| Space group | $P2_12_12_1$ | $P2_12_12_1$ |
| Cell Dimensions (a,b,c) (Å) | 50.81, 81.24, 110.07 | 50.93, 81.55, 110.53 |
| Resolution (Å) | 45.56 – 1.85 (1.89 – 1.85) | 46.25 – 1.65 (1.68 – 1.65) |
| Total number of observations | 535056 (32820) | 761967 (35727) |
| Number of unique observations | 39747 (2437) | 56241 (2718) |
| Multiplicity | 13.5 (13.5) | 13.5 (13.1) |
| Data completeness (%) | 100 (100) | 100 (100) |
| I/σ<sub>I</sub> | 9.2 (2.6) | 12.0 (2.4) |
| Mn(I) half-set correlation CC<sub>(1/2)</sub> | 0.996 (0.839) | 0.998 (0.728) |
| R<sub>pim</sub><sup>a</sup> (%) | 6.1 (62.7) | 4.8 (67.9) |
| Refinement Statistics |  |  |
| Non-hydrogen atoms |  |  |
| Protein | 3229 | 3232 |
| Water | 329 | 510 |
| R<sub>factor</sub><sup>b</sup> (%) | 17.8 | 18.3 |
| R<sub>free</sub><sup>b</sup> (%) | 22.3 | 22.0 |
| Rms deviations from ideality |  |  |
| Bond lengths (Å) | 0.011 | 0.005 |
| Bond angles (°) | 1.484 | 0.994 |
| Ramachandran plot (%) |  |  |
| Allowed region | 99.75 | 99.75 |
| Disallowed region | 0.25 | 0.25 |
| PDB code | 8ELH | 8ELG |

## Results

### Citizen Science cohort population characteristics

Our study population was overwhelmingly comprised of individuals with mild disease, with only two individuals reporting hospitalization for COVID-19. Our final cohort consisted of 1,428 individuals who reported a positive test for active SARS-CoV-2 infection and self-identified as White. Among these respondents, 136 (9.5%) reported having remained asymptomatic for at least two weeks before and after a positive test for virus. Median age was increased in asymptomatic compared to symptomatic individuals (asymptomatic=41, symptomatic=33, *p* < 0.001), and we observed increased reporting for asymptomatic infection in males (*p*=0.02). Basic demographics for all subjects are given in **Table 3**.

**Table 3.** Study population demographics.

| **Total** | **Symptomatic** | **Asymptomatic** |  |
| --- | --- | --- | --- |
| 1428 | 1292 | 136 |  |
|  | 90.5% | 9.5% |  |
|  | **Male** | **Female** | **Age**<br>**(median)** |
| **All** | 19% | 81% | 34 |
| **Symptomatic** | 18% | 82% | 33 |
| **Asymptomatic** | 26% | 74% | 41 |

$p$ = p-value

We also collected data on several diseases and conditions that might impact COVID-19 disease course. Among all individuals who reported a positive test for virus (n=1,428), 67% reported no known COVID-19-associated comorbidities. The full list of reported diseases and conditions and their frequency in this cohort is given **Supplementary Table S6**.

### HLA-B*15:01 is associated with asymptomatic SARS-CoV-2 infection

We aimed to identify whether HLA variation impacts the likelihood that an individual will remain asymptomatic after SARS-CoV-2 infection. We analyzed high-resolution genotyping for five highly polymorphic *HLA* class I and class II genes (*HLA-A, HLA-B, HLA-C, HLA-DRB1, HLA-DQB1*) in our discovery (Citizen Science) cohort. Data analysis included the first two fields of the allele name as described in the HLA nomenclature, representing the complete molecule at polypeptide sequence resolution.

We found the allele *HLA-B*15:01* was significantly overrepresented in asymptomatic individuals relative to symptomatic individuals (frequency =0.1103 vs. 0.0495; OR=2.38; 95%CI 1.51-3.65, *p* = 3 × 10^−5^, *p*^corr^ = 0.002). No other *HLA* allele at any locus was found to be significantly associated after correction for multiple comparisons. Allele frequencies for all loci are given in **Supplementary Table S7**.

To adjust for the effect of comorbid conditions, as well as sex and age differences in asymptomatic vs. symptomatic patients, we fitted a series of regression models but did not find any impact of patient-reported comorbidities on the likelihood of asymptomatic disease. Thus, our final model adjusted only for age and sex, which again showed a significant association of *HLA-B*15:01* with asymptomatic infection after adjustment for these variables (OR=2.40 95% CI = 1.54-3.64; *p* = 5.67 ×10^−5^, *p*^corr^ = 0.003, **Table 4**).

**Table 4.**
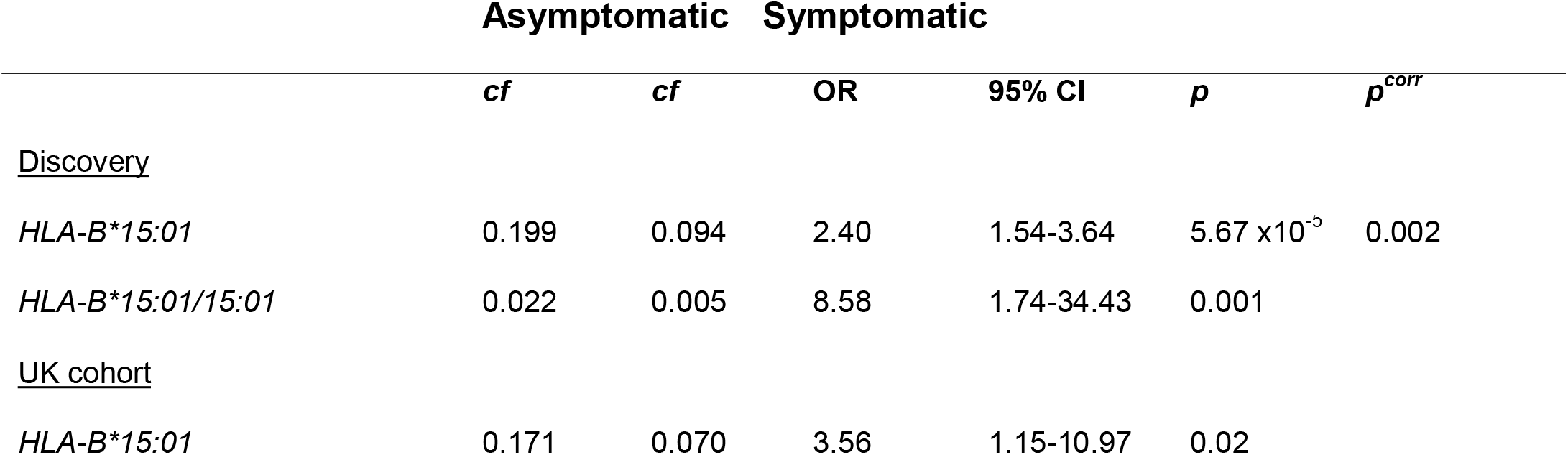
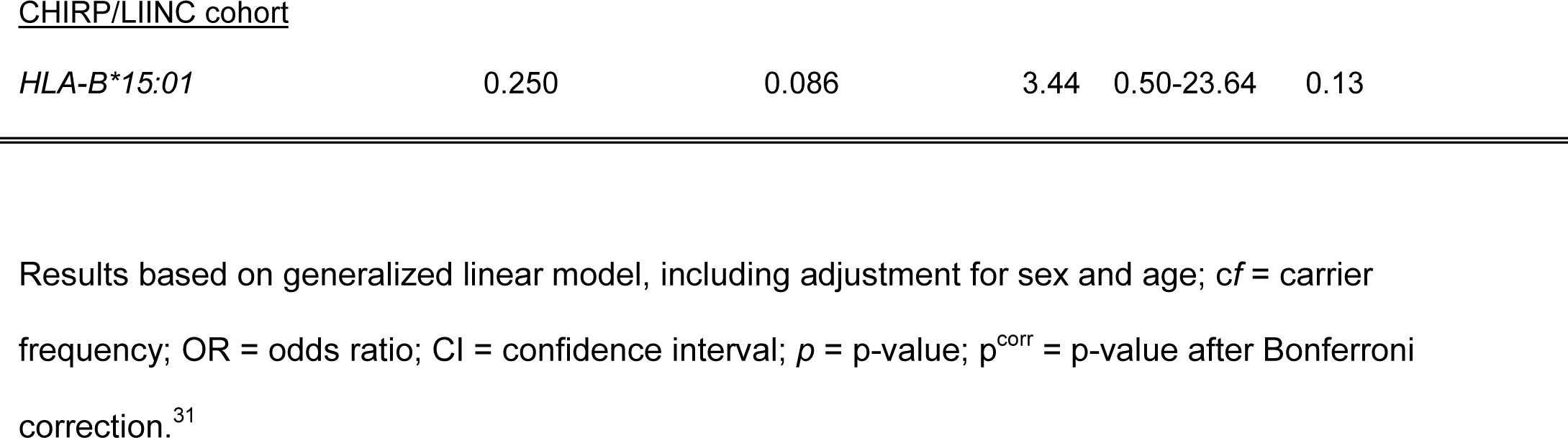
HLA-B*15:01 is associated with SARS-CoV-2 asymptomatic infection.

Finally, we observed a strong additive effect for the associated genotype. Individuals who carry two copies of *HLA-B*15:01* are more than eight times more likely to remain asymptomatic than individuals carrying other genotypes (OR = 8.58, 95%CI = 1.74-34.43, *p* = 0.001). Overall, one in five individuals (20%) who remained asymptomatic after infection carried *HLA-B*15:01*, compared to 9% among patients reporting symptoms.

### HLA-B*15:01 association with asymptomatic SARS-CoV-2 infection is enhanced by the presence of HLA-DRB1*04:01

To understand whether additional *HLA* alleles might interact with *HLA-B*15:01* in asymptomatic infection, we tested all pairwise two-locus haplotypes containing *HLA-B*. Overall, haplotypic associations for *HLA-B∼HLA-DRB1* and *HLA-A∼HLA-B* were found to be significant at *p*=0.01. Examining specific allelic haplotypes, these associations were driven by two *HLA-B*15:01* haplotypes: *HLA-B*15:01∼HLA-DRB1*04:01*, and *HLA-A*02:01∼HLA-B*15:01* (**Supplementary Tables S8-9**).

After adjusting for sex and age, only the combination of *HLA-B*15:01* and *HLA-DRB1*04:01* remained significant after correction for multiple comparisons (*p* = 3 × 10^−4^, *p*^corr^ = 0.01). We found an odds ratio for this combination (OR=3.17, 95% CI=1.65-5.80) that exceeds that for *HLA-B*15:01* alone, suggesting that while not significantly associated with the asymptomatic infection on its own in this cohort, the class II allele *HLA-DRB1*04:01* enhances the effect of *HLA-B*15:01*.

### The HLA-B*15:01 association with asymptomatic SARS-CoV-2 infection is replicated in two independent cohorts

To confirm our finding establishing the *HLA-B*15:01* association with asymptomatic infection in SARS-CoV-2, we examined two independent cohorts of patients with European ancestry. We first undertook a re-analysis of the primary *HLA* genotype data in a U.K. cohort previously reported in Langton et al., which had not directly tested *HLA-B*15:01* with respect to asymptomatic infection. Testing only the allele of interest, we found that *HLA-B*15:01* is strongly associated with asymptomatic infection in this cohort when adjusting for sex and age (*p*=0.02; OR=3.56, 95% CI=1.15-10.94). Similar to our discovery cohort, we found that the carrier frequency for *HLA-B*15:01* was 17% in asymptomatic subjects, compared to 7% in symptomatic patients (**Table 4**).

We next examined the association of *HLA-B*15:01* with asymptomatic infection in the combined UCSF prospective longitudinal COVID-19 Host Immune Response Pathogenesis (CHIRP) and Long-term Impact of Infection with Novel Coronavirus (LIINC) cohorts. Here 12/82 subjects with European ancestry were identified as having an asymptomatic disease course. We found again that carrier frequency of *HLA-B*15:01* was exceptionally high (25%) in asymptomatic subjects compared to symptomatic patients (8.6%). While power was somewhat limited by sample size, the findings are strongly trending in support of our finding of a strong association of this allele with asymptomatic disease (*p*=0.13; OR=3.44, 95% CI=0.50-23.64; **Table 4**). Finally, a meta-analysis across all three datasets (Citizen Science, UK, CHIRP/LIINC) confirmed the strong and consistent association of *HLA-B*15:01* with asymptomatic infection (p<10^−4^; OR=2.55, 95% CI=1.73-3.77, **Supplementary Figure S1**).

### HLA-B*15:01-restricted T cells from healthy (pre-pandemic) donors are reactive to SARS-CoV-2 and display a memory phenotype

HLA tetramers (pHLA) consist of four HLA molecules associated with a specific peptide and bound to a fluorochrome. Due to their high avidity for their cognate T-cell receptors, they have been systematically used to visualize and quantify low frequency antigen-specific T cells *ex vivo* using flow cytometry.^44,45^ We focused on four SARS-CoV-2 epitopes (CVADYSVLY, HVGEIPVAY, NQKLIANQF, and RVAGDSGFAAY) previously shown to elicit cellular immunity mediated by cytotoxic CD8+ T cells in COVID-19 patients carrying HLA-B*15:01.^46–50^ Next, we performed *ex vivo* pMHC tetramer evaluation with the SARS-CoV-2 peptides to detect antigen-specific CD8+ T cells in nine pre-pandemic PBMC samples. We observed tetramer-positive CD8+ T cells for three of the SARS-CoV-2 epitopes (**Figure 3A-B**). NQKLIANQF was detectable in the highest proportion of samples (55.6%) with a mean frequency of 1.1 × 10^−3^. Remarkably, 100% of NQKLIANQF tetramer-positive CD8+ T cells were memory T cells, indicating pre-existent T-cell immunity against SARS-CoV-2 in individuals carrying HLA-B*15:01 who did not have any previous contact with the virus (**Figure 3C**).

**Figure 3.**
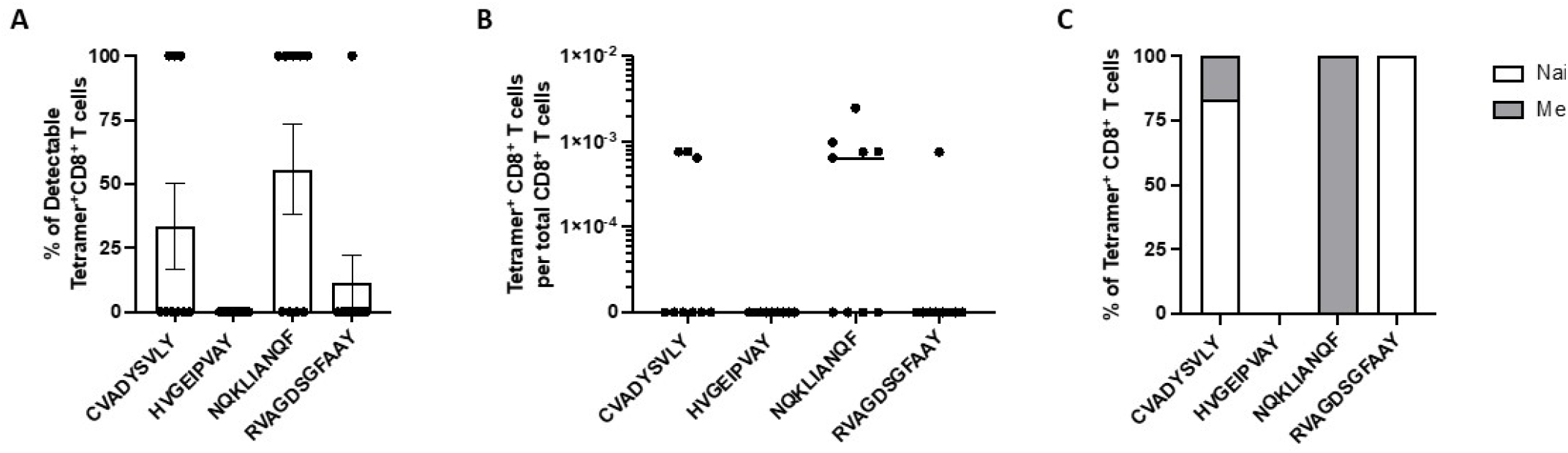
T cell reactivity in pre-pandemic samples from individuals carrying HLA-B*15:01. *Ex vivo* tetramer analysis for the four indicated epitopes was performed in nine pre-pandemic donor samples. The proportion of detectable tetramer-positive CD8+ T cells (A) and their frequencies are shown (B). The proportion of naïve (CD45RA+ CCR7+) and memory (combination of CD45RA-CCR7-, CD45RA-CCR7+, CD45RA+ CCR7-) tetramer-positive CD8+ T cells is shown in C.

### Peptides from SARS-CoV-2, OC43-CoV, and HKU1-CoV adopt the same conformation in the HLA-B*15:01 molecule

The HLA-B*15:01-restricted peptide NQKLIANQF is conserved among all SARS-CoV-2 variants and differs by only one amino acid position from HKU1-CoV and OC43-CoV peptides (**Supplementary Figure S2**). Importantly, T cells from SARS-CoV-2 infected *HLA-B*15:01* positive patients reactive to NQKLIANQF had previously been shown by Minervina et al. (2022)^50^ to be cross-reactive to the related peptide from seasonal coronaviruses. Because our *ex vivo* analysis of pHLA furthered these results by suggesting pre-existing T-cell immunity in pre-pandemic samples, we sought to investigate whether the amino acid change between the NQK peptides from SARS-CoV-2 and the seasonal coronaviruses HKU1-CoV and OC43-CoV impact the stability of HLA-B*15:01.

We refolded the HLA-B*15:01 molecule in the presence of each peptide (NQKLIAN**Q**F and NQKLIAN**A**F), and undertook thermal melt assay using Differential Scanning Fluorimetry (DSF). Both pHLA complexes exhibited the same thermal melt point (Tm, **Table 1, Figure 4A**), indicating that the Q>A amino acid change does not impact on the overall stability of the pHLA.

**Figure 4.**
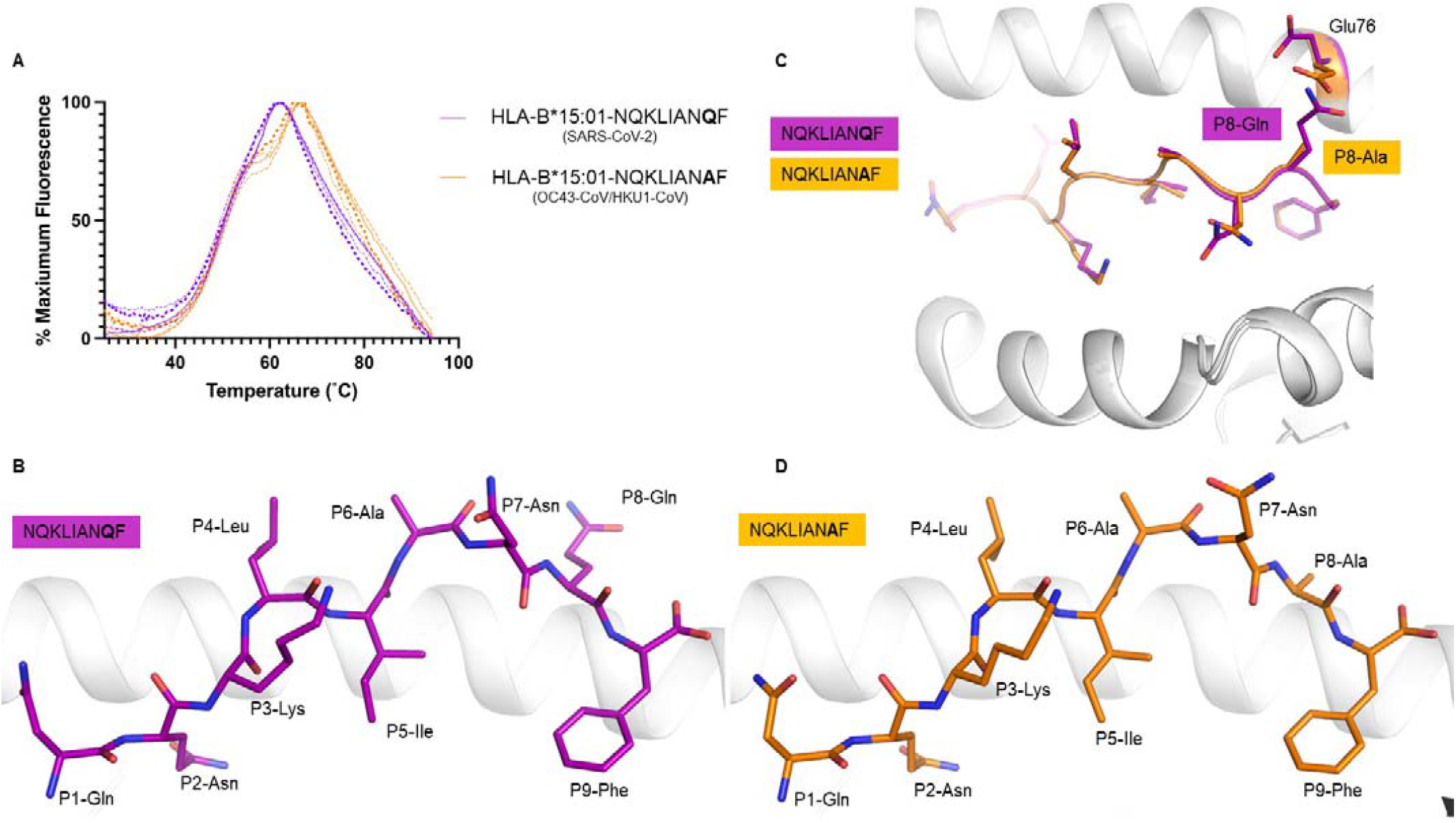
HLA-B*15:01 interaction with peptides derived from SARS-CoV-2 and seasonal coronaviruses. (**A**) DSF plots show normalized fluorescence intensity versus temperature for HLA-B*15:01 in complex with the NQKLIAN**Q**F (SARS-CoV-2) peptide (purple) or NQKLIAN**A**F (HKU1-CoV and OC43-CoV) peptide (orange) measured at 5 and 10 μM concentrations (n=2). (**B**) Crystal structure of HLA-B*15:01-NQKLIAN**Q**F with the HLA-B*15:01 in white cartoon and the peptide in purple stick (**C**) superposition of the structures of HLA-B*15:01-peptides (**D**) structure of HLA-B*15:01-NQKLIAN**A**F with the HLA-B*15:01 in white cartoon and the peptide in orange stick.

Next, we inquired if this amino acid substitution could affect the peptide presentation by HLA-B*15:01. We crystallized each peptide in complex with the HLA-B*15:01 and solved their structures at high resolution (**Table 2, Figure 4B-D, Supplementary Figure S3 A-D**). Overall, the NQKLIAN**Q**F (SARS-CoV-2) adopted a canonical conformation within the antigen binding cleft of the HLA-B*15:01 molecule.^51^ The Gln at position 2 (P2) was deeply inserted into the HLA-B*15:01 B pocket, while the P9-Phe acted as another primary anchor residue within the F pocket. The central part of the peptide was more mobile than the rest of the peptide, with weaker electron density from P4-Leu to P6-Ala as shown by the higher B factor of their backbone (**Supplementary Figure S3 E**). The peptide NQKLIAN**Q**F exposed to the solvent, and potentially to circulating T cells, five of its nine residues (P1-Asn, P4-Leu, P6-Ala, P5-Asn, and P8-Gln).

The peptide NQKLIAN**A**F (seasonal coronaviruses HKU1-CoV and OC43-CoV) bound in a similar fashion into the HLA-B*15:01 cleft (**Figure 4C-D, Supplementary Figure S3**), adopting a canonical extended conformation in the HLA-B*15:01 molecule. The central part of the NQKLIAN**A**F peptide was also mobile as shown by a weaker electron density than the rest of the peptide (**Supplementary Figure S3 C-D**). In addition, the same five residues from the peptide NQKLIAN**Q**F were solvent exposed in the NQKLIAN**A**F peptide, presenting the same surface, with the exception of the P8 position, to the T cells.

The superposition of the two pHLA structures revealed very little difference between the two complexes, with a root mean square deviation (r.m.s.d.) of 0.08 Å for the Cα atoms of the antigen binding cleft (residue 1 – 180) and of 0.12 Å for the peptides. Overall there were no structural changes between the two complexes, with limited impact of the sequence difference at P8 of the peptides onto the peptide presentation (**Figure 4C**). This also gives a rational basis for the similar thermal stability observed for the two pHLA complexes (**Table 1**). While the P8 mutation did not impact the overall structure, there was some local rearrangement around this residue. The P8 residue was solvent exposed in both peptides, and the larger P8-Gln in the NQKLIAN**Q**F peptide provoked a shift of the Glu76 side chain to avoid steric clashes.

## Discussion

Understanding the biological underpinning of asymptomatic infection with SARS-CoV-2 has important implications for public health measures, vaccine design, and therapeutic development. Here we provide the first evidence of a genetic basis and the mechanistic explanation underlying asymptomatic disease. Leveraging a large database and mobile technology in this crowd-sourced study, we reveal important insight into the immunogenetic underpinnings of asymptomatic SARS-CoV-2 infection. Our innovative use of a mobile application and a pre-existing database for medical research allowed us to screen nearly 30,000 individuals previously genotyped for *HLA* for viral infection and disease course. We augment our findings of a strong HLA association with asymptomatic disease course in this unique cohort with functional and structural studies to support a model of pre-existing immunity to explain the observed *HLA* association.

We show that among participants reporting a positive test result for SARS-CoV-2, *HLA-B*15:01* is significantly associated with asymptomatic infection. We observed that individuals carrying this common allele (approximately 10% in individuals with European ancestry) are more than twice as likely to remain asymptomatic after SARS-CoV-2 infection than those who do not, and an astonishing effect for *HLA-B*15:01* homozygosity increasing more than eight times the chance of remaining asymptomatic. This observation suggests important features of early infection with SARS-CoV-2. Supporting the role of *HLA-B*15:01* in mediating asymptomatic infection, these results replicated in two independent data sets.

Despite a growing number of published studies, the role of *HLA* variation in COVID-19 has remained unclear, with no clear consensus in the literature to-date, and, notably, few studies examining asymptomatic infection as a primary phenotype^26^. Our re-analysis of the primary data underlying a reported association of *HLA-DRB1*04:01* with asymptomatic infection^27^ uncovered clear evidence for the role of *HLA-B*15:01* in asymptomatic disease, which had not been examined in the initial study. While the data in our discovery cohort did not corroborate the association for *HLA-DRB1*04:01* alone, we did find that this allele enhanced the effect of *HLA-B*15:01* when the pair were in combination. We note that this is the *HLA-DRB1* allele most commonly associated with *HLA-B*15:01* in individuals in the U.S. who self-identify as White,^28^ and thus it is difficult to differentiate a real effect from one related to linkage disequilibrium between these loci unless directly tested. Likewise, another recent paper describing an association of *HLA-DRB1* alleles with asymptomatic infection did not genotype for *HLA-B*.^52^ Finally, two other large studies that employed patient questionnaires regarding symptoms did not consider the most mild symptoms common in SARS-CoV-2 infection (e.g. “runny nose” and “scratchy throat”), resulting in a much less stringent definition of asymptomatic infection than we considered here.^53,54^

For decades, respiratory tract infections have been a major public health concern, representing a significant burden, particularly for young children and the elderly.^55–57^ Four strains of seasonal coronaviruses (229E-CoV, NL63-CoV, OC43-CoV, and HKU1-CoV) represent 15% to 30% of all respiratory tract infections every year.^58,59^ Interestingly, previous work has shown that certain SARS-CoV-2 peptides cross-react with seasonal coronaviruses indicating that long-lasting protective immunity due to T-cell cross-reactivity can potentially limit the severity of COVID-19.^47^ In addition, Minervina et al.^50^ have recently demonstrated T-cell cross reactivity for SARS-CoV-2 and seasonal coronaviruses for a HLA-B*15:01-restricted immunodominant epitope (NQKLIAN**Q**F) in individuals who received two doses of the Pfizer-BioNTech BNT162b2 mRNA vaccine. To test the hypothesis that HLA-B*15:01 can mediate asymptomatic disease via pre-existing T-cell immunity, we analyzed immunodominant epitopes in T cells from human PBMCs from pre-pandemic healthy individuals. We observed that T cells from more than half of pre-pandemic healthy donors carrying HLA-B*15:01 were reactive to the SARS-CoV-2 peptide NQKLIAN**Q**F, and 100% of the reactive cells displayed memory phenotype. The sequence identity between SARS-CoV-2 peptides and seasonal coronaviruses, except for a single amino acid substitution, could explain the T-cell cross-reactivity. However, a direct demonstration that peptides from SARS-CoV-2 and the seasonal coronaviruses OC43-CoV and HKU1-CoV are stable into the HLA-B*15:01 cleft was necessary to further corroborate our hypothesis.

Through our examination of the crystal structures of the HLA-B*15:01 molecule in the presence of each peptide we demonstrated that both NQKLIAN**Q**F (SARS-CoV-2) and NQKLIAN**A**F (OC43-CoV, and HKU1-CoV) spike peptides share similar ability to stabilize the HLA-B*15:01 molecule, and are presented in a similar conformation from the HLA-B*15:01, providing the molecular basis for T-cell cross-reactivity and pre-existing immunity. This observation is in accordance with previous work in uninfected HLA-B*07:02^+^ individuals able to recognize the N_105-113_ peptide derived from SARS-CoV-2 due to the presence of cross-reactive T cells recognizing the homologous N_105-113_ peptide from OC43-CoV and HKU1-CoV.^47^ Interestingly, this T-cell cross reactivity has been associated with less severe COVID-19 disease.^60^

Examination of T cells in patients with asymptomatic SARS-CoV-2 infection has suggested similarly robust T cell responses in these patients compared to those with symptomatic disease.^61^ In addition, the finding that T cells in asymptomatic infection secrete higher quantities of IFN-γ than those in symptomatic patients early in infection^61,62^ supports a role for memory T cells at this stage.^63^ While the current literature is mixed regarding cross-reactive CD8+ T cells specific to SARS-CoV-2, this might be explained by HLA specificity.^64,65^ The fact that asymptomatic individuals in our study were less likely than symptomatic subjects to report a positive test for antibodies against SARS-CoV-2 (**Supplementary Table S10**) suggests a strong early anti-viral response and may corroborate the critical role of CD8+ T cytotoxicity mediated by HLA class I in eliminating SARS-CoV-2 infected cells. Altogether, our results strongly support the hypothesis that HLA-B*15:01 mediates asymptomatic COVID-19 disease via pre-existing T-cell immunity due to previous exposure to HKU1-CoV and OC43-CoV.

One limitation of this study is that all testing results and symptoms in our discovery cohort are self-reported. We recognize that this may result in some margin of error in our results. However, we have previously validated this approach by verifying test results in a subset of the participants.^66^ Additionally, we find a remarkably consistent genetic association across the study and in two independent cohorts where asymptomatic disease was clinician-defined, pointing to a true biological feature. Another limitation is that we only tested four SARS-CoV-2 peptides in our *ex vivo* analysis. The search for additional candidate peptides will be facilitated as more studies analyze T-cell reactivity in patients carrying HLA-B*15:01, similarly to that performed by Minervina et al.^50^ However, despite this limitation, we identified at least one SARS-CoV-2 peptide previously known to be immunodominant in SARS-CoV-2 infection that was reactive to memory T-cells from HLA-B*15:01+ individuals collected before the pandemic.

In summary, we have demonstrated a strong and significant association of a common *HLA* class I allele, *HLA-B*15:01*, with asymptomatic infection with SARS-CoV-2. We demonstrated that B*15:01+ T cells from pre-pandemic samples were reactive to an immunodominant SARS-CoV-2 peptide that shares high sequence similarity with peptides from seasonal coronaviruses. We provided the molecular basis of cross-reactivity by showing HLA-B*15:01 has the ability to stabilize and present peptides from HKU1-CoV and OC43-CoV similarly to the immunodominant peptide from SARS-CoV-2. Our results have important implications for understanding early infection and the mechanism underlying early viral clearance and may lay the groundwork for refinement of vaccine development and therapeutic options in early disease.

## Data Availability

All data is provided in the manuscript and supplementary data

## Acknowledgment

This study was funded by grants R01AI159260 and 3U2CEB021881-05S1 from the National Institutes of Health, and the National Health and Medical Research Council (NHMRC) and Medical Research Future Fund (MRFF), NHMRC SRF (#1159272). The CIBMTR is supported primarily by Public Health Service U24CA076518 from the National Cancer Institute (NCI), the National Heart, Lung and Blood Institute (NHLBI) and the National Institute of Allergy and Infectious Diseases (NIAID); HHSH250201700006C from the Health Resources and Services Administration (HRSA); and N00014-20-1-2832 and N00014-21-1-2954 from the Office of Naval Research; Support is also provided by Be the Match Foundation, the Medical College of Wisconsin and the National Marrow Donor Program. The views expressed in this article do not reflect the official policy or position of the National Institute of Health, the Department of the Navy, the Department of Defense, Health Resources and Services Administration (HRSA) or any other agency of the U.S. Government. The authors wish to thank the thousands of volunteer bone marrow donors who took the time to participate in this study.

