## Supplemental Figures for "A common allele of HLA mediates asymptomatic SARS-CoV-2 infection"

### Supplementary Figures

**Supplementary Figure S1. Meta-analysis across discovery and replication cohorts.**

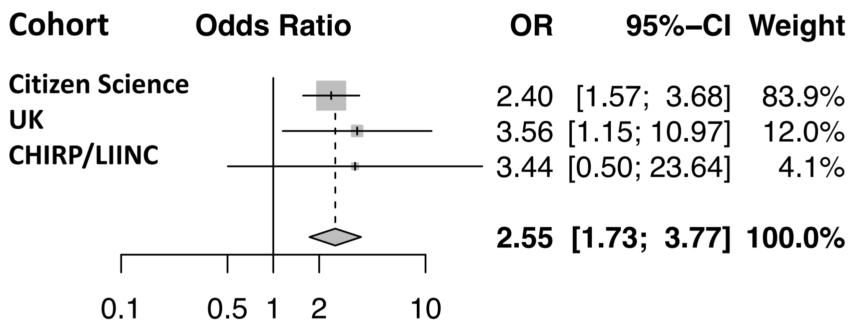


Meta-analysis p-value < $10^{-4}$. OR = odds ratio; CI – confidence interval

**Supplementary Figure S2. Partial amino acid alignment among coronaviruses spike protein**

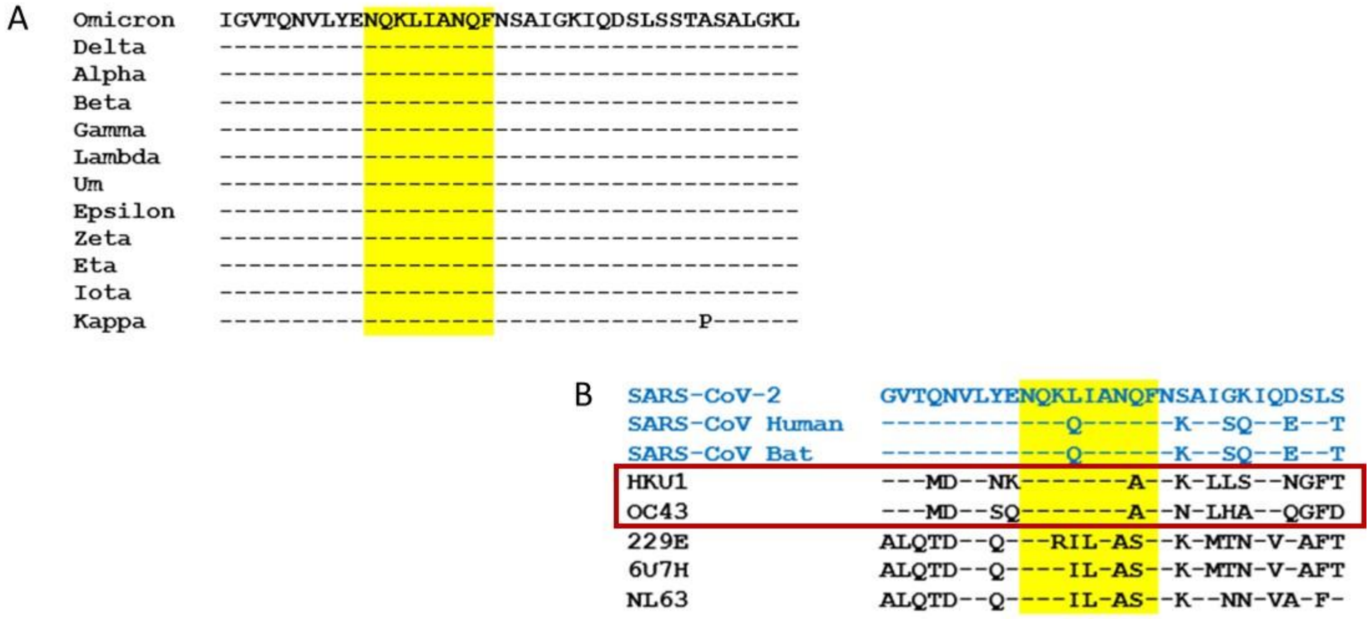


The yellow shade highlights the SARS-CoV-2 peptide NQKLIANQF as the reference sequence. A) Alignment of the NQKLIANQF across SARS-CoV-2 variants. B) Alignment of NQKLIANQF across multiple coronaviruses. Blue marks other SARS coronaviruses and the red rectangle marks two seasonal coronaviruses for which there is only one amino acid difference for the reference peptide NQKLIANQF.

**Supplementary Figure S3. Electron density map for the NQK peptides from SARS-CoV-2 and other coronaviruses.**

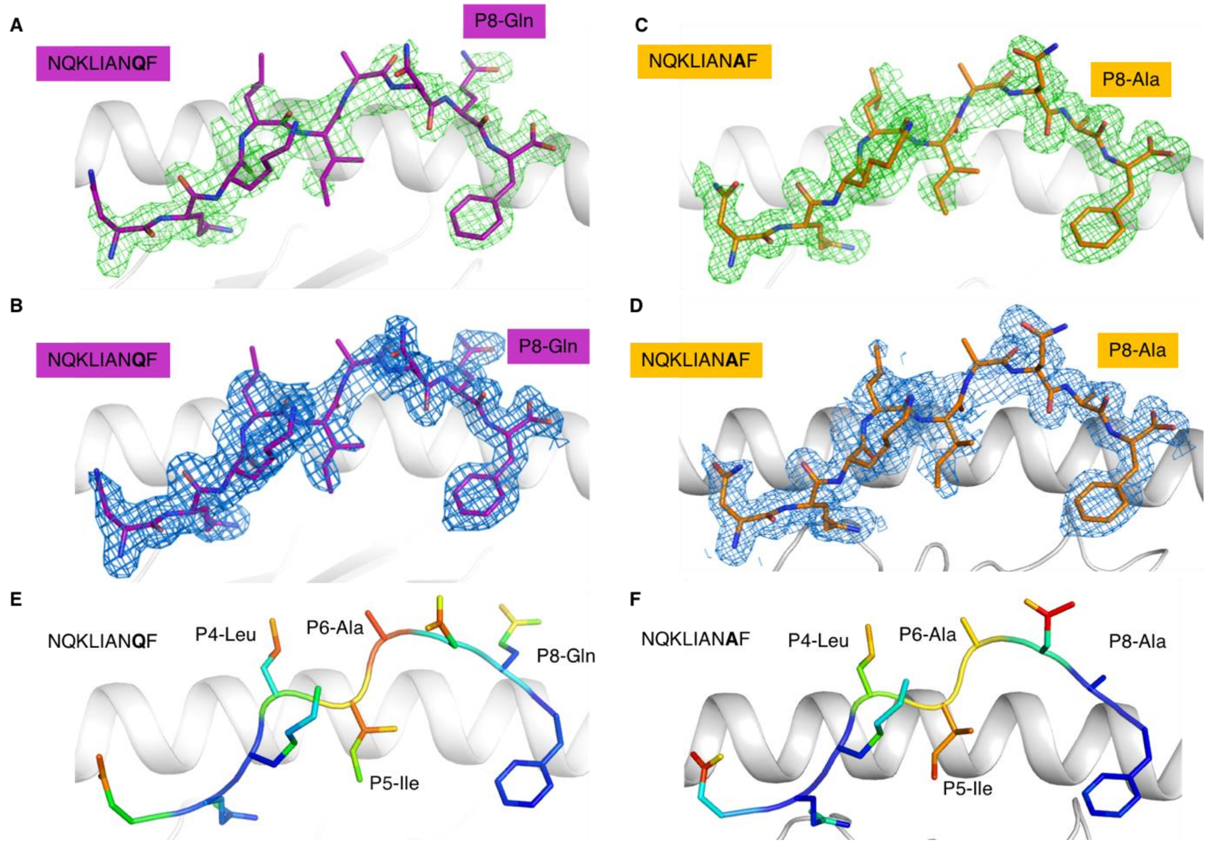


(**A**) and (**B**) Electron density map around the NQKLIAN**Q**F (SARS-CoV-2) peptide (purple stick) in blue at 1$\sigma$ for the 2Fo-Fc map and in green at 3$\sigma$ for the Fo-Fc map, respectively. (**C**) and (**D**) Electron density map around the NQKLIAN**A**F (HKU1-CoV and OC43-CoV) peptide (orange stick) in blue at 1$\sigma$ for the 2Fo-Fc map and in green at 3$\sigma$ for the Fo-Fc map, respectively. (**E**) and (**F**) $B_{factor}$ analysis of the atoms of the NQKLIAN**Q**F and NQKLIAN**A**F peptides, respectively. The atoms are colored accordingly to their $B_{factor}$ from low (blue) to high (red) that indicates the mobility of each atom.
